# Muscle microRNAs in the cerebrospinal fluid predict clinical response to nusinersen therapy in type II and type III spinal muscular atrophy patients

**DOI:** 10.1101/2021.07.29.21261322

**Authors:** Iddo Magen, Sharon Aharoni, Nancy Sarah Yacovzada, Itay Tokatly Latzer, Christiano R R Alves, Liora Sagi, Aviva Fattal-Valevski, Kathryn J Swoboda, Jacob Katz, Elchanan Bruckheimer, Yoram Nevo, Eran Hornstein

## Abstract

**Objective:** The antisense oligonucleotide nusinersen (spinraza) regulates splicing of the *survival motor neuron 2* (*SMN2)* messenger RNA to increase SMN protein expression and has improved ventilator free survival and motor function outcomes in infantile onset forms of SMA, treated early in the course of the disease. However, the response in later onset forms of SMA is highly variable and dependent on symptom severity and disease duration at treatment initiation. Therefore, we aimed to identify novel noninvasive biomarkers that could predict the response to nusinersen in type II and III SMA patients.

**Methods:** 34 SMA patients were included. We applied next-generation sequencing to identify microRNAs in the cerebrospinal fluid (CSF) as candidate biomarkers predicting response to nusinersen. Hammersmith Functional Motor Scale Expanded (HFMSE), was conducted at baseline and 6 months post initiation of nusinersen therapy to assess motor function. Patients changing by ≥ 3 or ≤0 points in the HFMSE total score were considered as responders or non-responders, respectively.

**Results:** Lower baseline levels of two muscle microRNAs (miR-206 and miR-133), alone or in combination, predicted the pre-determined clinical response to nusinersen after 6 months therapy. Moreover, miR-206 levels were inversely correlated with the HFMSE score.

**Conclusions:** Lower miR-206 and miR-133 in the CSF predict more robust clinical response to nusinersen treatment in later onset SMA patients. These novel findings have high clinical relevance for identifying early treatment response to nusinsersen in later onset SMA patients and call to test the ability of miRNAs to predict more sustained long-term benefit.

## Introduction

Spinal muscular atrophy (SMA) is a genetic disease with an incidence of ∼ 1/11,000 live births, and a carrier frequency of ∼1/40 (1, 2). SMA is characterized by muscle weakness and atrophy, resulting from progressive degeneration of lower motor neurons in the spinal cord and the brain stem nuclei. Historically, SMA subtype classification is based on age of onset and the maximum motor abilities achieved. SMA type I patients have onset in early infancy and never sit. SMA type II patients have later infantile onset but never walk. SMA type III patients have more variable childhood onset, and achieve the ability to walk (3). SMA IV patients typically present with milder muscle weakness in the second or third decade of life (4-7). The cause of SMA are homozygous deletion or compound heterozygous mutation involving exon 7 of the Survival Motor Neuron 1 (*SMN1*) gene. However, *SMN1* has a paralogous gene named *SMN2*, which undergoes alternative splicing, including the removal of exon 7, and produces ∼10% functional SMN protein (8, 9). Therefore, the number of *SMN2* copies correlates with phenotypic severity and is the main genetic disease modifier (10-12).

Nusinersen was the first drug approved by the US Food and Drug Administration (FDA) to precisely treat SMA. The drug is a synthetic antisense oligonucleotide that modulates pre-messenger RNA splicing of the *SMN2* gene. Despite the advance of novel molecular and gene therapies for SMA, nusinersen remains to this day the most widely used and available SMA disease-modifying therapy. Nusinersen has been proven efficient in young type I SMA patients, and it is dramatically changing the natural history of this disease (13, 14). However, the response in subjects with later-onset forms of SMA (type II and III) is more variable, with 30-40% presenting clinically meaningful improvement (15, 16). Therefore, better understanding of the variable treatment response in this population is important and novel biomarkers to predict treatment response to nusinersen in type II or type III SMA subjects are critical.

A main interest of our group has been to investigate the profile of microRNAs (miRNAs) in the disease context, since several endogenous non-coding RNAs are highly expressed in both neuronal and muscular tissues. Previous evidence indicates that some miRNAs are essential for motor neuron survival, and low expression levels have been demonstrated in motor neurons from an SMA mouse model and in postmortem neuronal tissues from patients with another motor neuron disease, amyotrophic lateral sclerosis (ALS) (17-19).

Moreover, miRNAs miR-1/133a/133b/206, are mainly expressed in the skeletal muscle, where they are thought to play an important role in myoblast proliferation and differentiation (20, 21). These are also known as myomiRs. Recent studies have demonstrated that the expression of myomiRs can be detected in biofluids from ALS patients (22-24), suggesting a potential role of these molecules in early diagnosis of the disease. In addition, serum myomiRs are in correlation with the response to ongoing therapy in SMA type II and III (25). However, a prospective study that identifies responders to Nusinersen therapy, based on a basal, pre-treatment, molecular profile, was not yet reported.

On the basis of understanding the miRNA role in motor neuron diseases (17-19) and potential discovery of novel biomarkers (26-28), we sought to test the utility of specific miRNAs in the cerebrospinal fluid (CSF) as candidate molecules to predict a positive clinical response to nusinersen in type II/III SMA patients. We applied for the first time an unbiased next-generation sequencing to investigate the potential of cell-free microRNAs in the CSF of SMA patients receiving nusinersen therapy. Our novel findings indicate that the muscle miRNAs miR-206 and miR-133 predict the response to nusinersen treatment and, therefore, have the potential to help predict, when combined with other indicators, whether or not a given SMA patient is more or less likely to show an early response.

## Materials and Methods

### Subjects, Ethics and Motor Function

This study includes a cohort of 45 type II or type III SMA patients, that were recruited between November 2016 and July 2019 in three different medical centers: Schneider Children’s Medical Center of Israel (Israel, n=13), Dana-Dwek Children’s Hospital, Tel Aviv Medical Center (Israel, n=20) and Massachusetts General Hospital (MA, US, n=12). Approval for the study was provided by local ethical committees (Schneider Medical Center: RMC-0060-18; Tel Aviv Medical Center: 0347-18-TLV; Massachusetts General Hospital MGH #2016P000469), and it was conducted in accordance with the International Conference on Harmonization guidelines for Good Clinical Practice and the World Medical Association Declaration of Helsinki. Written informed parental consent was obtained from all participants and was the only inclusion criterion for treatment initiation. Nusinersen was administered on days 1, 15, 29, and 64 for the loading phase, followed by an additional maintenance dose after 4 additional months from the loading phase. Baseline demographic and clinical data were collected. Motor function was assessed at each visit by a physical therapist with SMA clinical trial training (29) using Hammersmith Functional Motor Scale Expanded (HFMSE) (30). Scoliosis was evaluated on every visit by physical examination and through spine / chest radiograph. Bilevel Positive Airway Pressure (BIPAP) machine usage data and subjective improvement in energy were collected from the medical records.

These assessments were all performed prior to RNA sequencing, and therefore the clinical assessor was oblivious to patients’ miRNA profile. CSF samples were collected on day 1 and 183 right before the nusinersen injection in each day and were stored frozen at −80°C.

### Inclusion and exclusion criteria

All 45 patients had a genetic diagnosis for SMA, and were followed up to this point. Inclusion criteria required non-hemolyzed CSF and full documentation of HFMSE score. Therefore, all type II/III SMA patients who signed informed consent for downstream analysis were enrolled without any pre-selection. Thus, the cohort was unbiased by any selective/representative inclusion/exclusion criteria

An improvement at the HFMSE final score ≥ 3 points was considered clinically significant (31, 32) and patients with such an improvement were considered as ‘responders’, while those with a change of HFMSE final score ≤0 were considered as ‘non-responders’. Samples from 11 patients with an intermediate clinical response to nusinersen (HFMSE improvement of 1 or 2 points) were excluded from main analyses, but are included in supplementary analyses (Figure S2, S5, S9, Table S2). Based on power analysis calculations, we found that this number was sufficient to obtain an effect size of 3 for change in miRNA level with a power of 95% and a p-value of 0.01.

### Small RNA Next Generation Sequencing

Total RNA was extracted from CSF using the miRNeasy micro kit (Qiagen, Hilden, Germany) and quantified with Qubit fluorometer using RNA broad range (BR) assay kit (Thermo Fisher Scientific, Waltham, MA). For small RNA next generation sequencing (RNA-seq), libraries were prepared from 7.5 ng of total RNA using the QIAseq miRNA Library Kit and QIAseq miRNA NGS 48 Index IL (Qiagen), by an experimenter who was blinded to the identity of patients from which the samples were collected, as well as to their HFMSE scores. Samples were randomly allocated to library preparation and sequencing in batches. Precise linear quantification of miRNA is achieved by using unique molecular identifiers (UMIs), of random 12-nucleotide after 3’ and 5’ adapter ligation, within the reverse transcription primers (26). cDNA libraries were amplified by PCR for 22 cycles, with a 3’ primer that includes a 6-nucleotide unique index, followed by on-bead size selection and cleaning. Library concentration was determined with Qubit fluorometer (dsDNA high sensitivity assay kit; Thermo Fisher Scientific, Waltham, MA) and library size with Tapestation D1000 (Agilent). Libraries with different indices were multiplexed and sequenced on NextSeq 500/550 v2 flow cell or Novaseq SP100 (Illumina), with 75bp single read and 6bp index read. Fastq files were de-multiplexed using the user-friendly transcriptome analysis pipeline (UTAP) (33). Human miRNAs, as defined by miRBase (34), were mapped using Geneglobe Data Analysis Center (Qiagen).

### Statistical analysis

As many as 2530 individual miRNA species were aligned to the human genome (GRCh37/hg19) across all samples. miRNAs with ≤100 UMIs on average across all samples were excluded from analysis. Sequencing data were normalized with DESeq2 package (35) under the assumption that miRNA counts followed negative binomial distribution and data were corrected for the library preparation batch in order to reduce its potential bias. Fold-change values in miRNA abundance between responders and non-responders were calculated as the ratio of normalized counts in responders (*i*.*e*. patients exhibiting meaningful clinical improvement) to the normalized counts in the non-responders (patients exhibiting lack of clinical improvement), and transformed to log base 2. Statistical significance was determined using the Wald test.

Feature scaling to the range of [0-1] was further done for DESeq2 normalized counts of miR-133a and miR-206 by applying min-max scaling, whereby the scaled value (x’) is calculated by subtracting the minimum value of each feature [min(x)] from each individual observation (x), and further divided by the difference between the maximum and the minimum value [max(x) – min(x)], as shown below

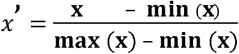

Scaled values were then summed to generate a devised feature termed miR-133a/206.

Correlation between miRNA levels and the clinical improvement in HFMSE score (HFMSE score post treatment – HFMSE score pretreatment) was calculated by both linear regression, wherein the improvement is a numerical value, and logistic regression, wherein the improvement is binarized into “failure” (HFMSE score change ≤ 0 between post and pretreatment) and “success” (HFMSE score change ≥ 3 between post and pretreatment). Logistic regression was performed with the aod and ggplot2 packages in R(36), either for a single explanatory variable (either one miRNA gene or a feature combined of two miRNAs) or as multinomial regression with multiple explanatory variables (a combination of different miRNAs or miRNAs and clinical features). The fitted logistic regression model is an exponential model from which predicted probabilities of “success” can be calculated based on values of the explanatory variable(s). Exponential equations for all models are available in **Tables S1 and S2**. Two values are derived from the logistic regression model: Chi-square (*χ*^2^), which expresses the goodness of fit for the whole model and Akaike’s information criterion (AIC)(37), an estimation of the model prediction error. Statistical significance was determined for Spearman rho and chi-square values, with p-values < 0.05 being considered statistically significant.

Leave-one-out-cross-validation was performed on the logistic regression model, whereby model learning was done in n-1 samples out of the total n, and tested on the leave-one-out sample, yielding a predicted success probability for each sample. This procedure was repeated n times. Receiver operating characteristic (ROC) curves were generated by plotting true positive rates (sensitivity), i.e. percentage of patients classified as responders, out of the total number of responders, against false positive rates (100% - specificity), i.e. percentage of patients classified as responders, out of the total number of non-responders, when each predicted success probability value is taken as a cut-off for binary classification. Area under the curve (AUC) and its respective P-value were calculated for ROC curves, given null hypothesis of AUC = 0.5. Graphs were generated with Prism 5 (GraphPad Software, San Diego, California, USA).

## Results

### Next-Generation Sequencing Analysis in the CSF of SMA Patients Treated with Nusinersen

We sought to explore miRNAs expressed in the CSF as potential biomarkers for monitoring response to nusinersen and to help determine, prior to treatment initiation, whether a patient was more or less likely to respond to the therapy. We used next generation sequencing to investigate, without an *a priori* bias, the comprehensive landscape of CSF miRNAs in 34 type II and type III SMA patients with documented demographic and clinical information available (**Table 1**). We analyzed RNA-seq data from CSF samples collected before treatment and analyzed these data in context of clinical response 6 months after initiation of nusinersen treatment (see study outline in **Figure 1)**. Data from additional 11 patients with HFMSE change of a single or two units (1,2) were excluded.

**Table 1.** Baseline characteristics at treatment onset: sex, SMA type, number of sitters/walkers, scoliosis, BIPAP use, age, disease duration from symptom onset and baseline HFMSE for each cohort. Data are presented for the total number of patients (45) and after exclusion of 11 patients (6 from Schneider, 3 from Tel Aviv and 2 from MGH) whose change in HFMSE score between after 6 months of treatment was 1 or 2. These patients were excluded from main analysis, but are included in supplementary analyses.

| Before exclusion |  |  |  |  |  |  |  |  |
| --- | --- | --- | --- | --- | --- | --- | --- | --- |
| Cohort | N<br>(% females) | Type<br>(II/III) | Sitters/<br>walkers | Scoliosis<br>(No/Yes) | BIPAP<br>use<br>(No/Yes) | Age in years,<br>median (range),<br>Number of adults | Disease duration<br>in years, median<br>(range) | Baseline HFMSE<br>median (range) |
| Schneider<br>Medical Center | 13 (69%) | 7/6 | 10/3 | 3/10 | 12/1 | 11.2 (2.3–19.3), 1 | 10.4<br>(1.5–16.7) | 26<br>(1–52) |
| Tel Aviv<br>Medical Center | 20 (55%) | 9/11 | 11/9 | 7/13 | 9/11 | 11.7 (1.7–24.4), 5 | 10.6<br>(0.2 - 21.8) | 11.5<br>(1–52) |
| Massachusetts<br>General<br>Hospital | 12 (42%) | 5/7 | 6/6 | 6/6 | 8/4 | 7.5 (2.1–56.6), 4 | 6.9<br>(1.9 - 48.6) | 41<br>(12 – 59) |
| Total | 45 (55%) | 21/24 | 27/18 | 16/29 | 29/16 | 11.0 (1.7- 6.6), 10 | 10.1<br>(0.2 – 48.6) | 28<br>(1 – 59) |
| After exclusion |  |  |  |  |  |  |  |  |
| Schneider<br>Medical Center | 7 (100%) | 4/3 | 5/2 | 2/5 | 0/7 | 6.4 (2.8–17.3), 0 | 4.4<br>(1.8 – 15.3) | 21<br>(3 – 52) |
| Tel Aviv<br>Medical Center | 17 (59%) | 8/9 | 8/9 | 7/10 | 8/9 | 11.1 (1.7–24.4), 3 | 10.1<br>(0.2 - 21.8) | 12<br>(2 – 50) |
| Massachusetts<br>General<br>Hospital | 10 (40%) | 4/6 | 5/5 | 5/5 | 7/3 | 11.1 (2.1–56.6), 4 | 9.1<br>(1.9 - 48.6) | 41<br>(12 – 59) |
| Total | 34 (53%) | 16/18 | 18/16 | 14/20 | 15/19 | 11.0 (1.7–56.6), 7 | 9.1<br>(0.2 – 48.6) | 27<br>(2 – 59) |

**Figure 1.**
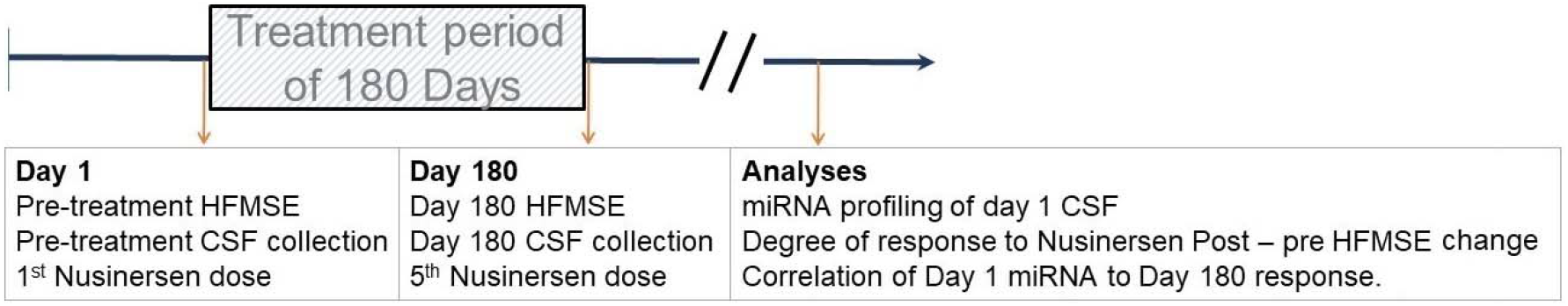
Summary of study outline. Patients were treated with nusinersen for 180 days. Behavioral assessment and miRNA quantification in CSF were conducted prior to treatment onset and prior to the last dose, and miRNA levels prior to treatment onset were compared between responders and non-responders as defined by HFMSE change post-treatment versus pre-treatment.

After next generation sequencing, 2530 miRNA were annotated to the genome. Only 68 miRNA species exceeded a cut-off of ≥100 unique molecular identifier (UMI) counts per sample, averaged on all samples. Five out of the 68 miRNAs changed post-treatment relative to pre-treatment with a fold change ≥1.2 and a p-value < 0.05 by Wald test (**Figure S1A**). Analysis of these patients by stratified response to therapy (≥ 3 or ≤0 HFMSE points for responders or non-responders, respectively), did not change the miRNA profile substantially (**Figure S1 B**,**C**).

These data suggest that only minor effects were observed on miRNA profile after 6 months of nusinersen therapy. Therefore, measurements of miRNAs in the CSF after treatment were not a main exploratory effort and did not significantly contribute to the study’s conclusions.

### Baseline miR-206 / miR-103 Levels Predict Beneficial Response to Nusinersen Therapy

The clinical response to nusinersen therapy is heterogeneous (38). We tested whether the miRNA profile at baseline would be different between patients that clinically responded to nusinersen therapy (an increase of ≥ 3 points in the HFMSE total score (32)) and those that poorly responded (HFMSE score change ≤ 0 points).

In responders’ CSF, two miRNAs differed in a significant manner relative to non-responders: miR-103b increased by 2.6-fold (Wald test: p = 0.002, **Figure 2A**), and miR-206 decreased by 1.8-fold (p = 0.048). Responders did not differ from non-responders in their clinical characteristics (**Figure 2B**). During these analyses, we noticed that similar to the downregulated miR-206 levels, other miRNAs that are known to be expressed mainly in the skeletal muscle (*i*.*e*. myomiRs) were relatively low in CSF of responders, including the miR-1-3p, miR-133a-3p and miR-133b, with 1.4-fold, 1.6-fold, and 2.2-fold, respectively. Our conclusion remains unchanged also when 11 additional patients with intermediate change in the HFMSE scores, which reflect modest response to therapy, were included as responders to therapy (**Figure S2**). We also note that miR-1180-5p and miR-6849 were downregulated at baseline in patients that self-reported energy improvement after therapy vs those who did not reporteimprovement (p<0.05, Wald test **Figure S3**). Therefore, miR-206 / miR-103 levels predict beneficial objective response to nusinersen therapy.

**Figure 2.**
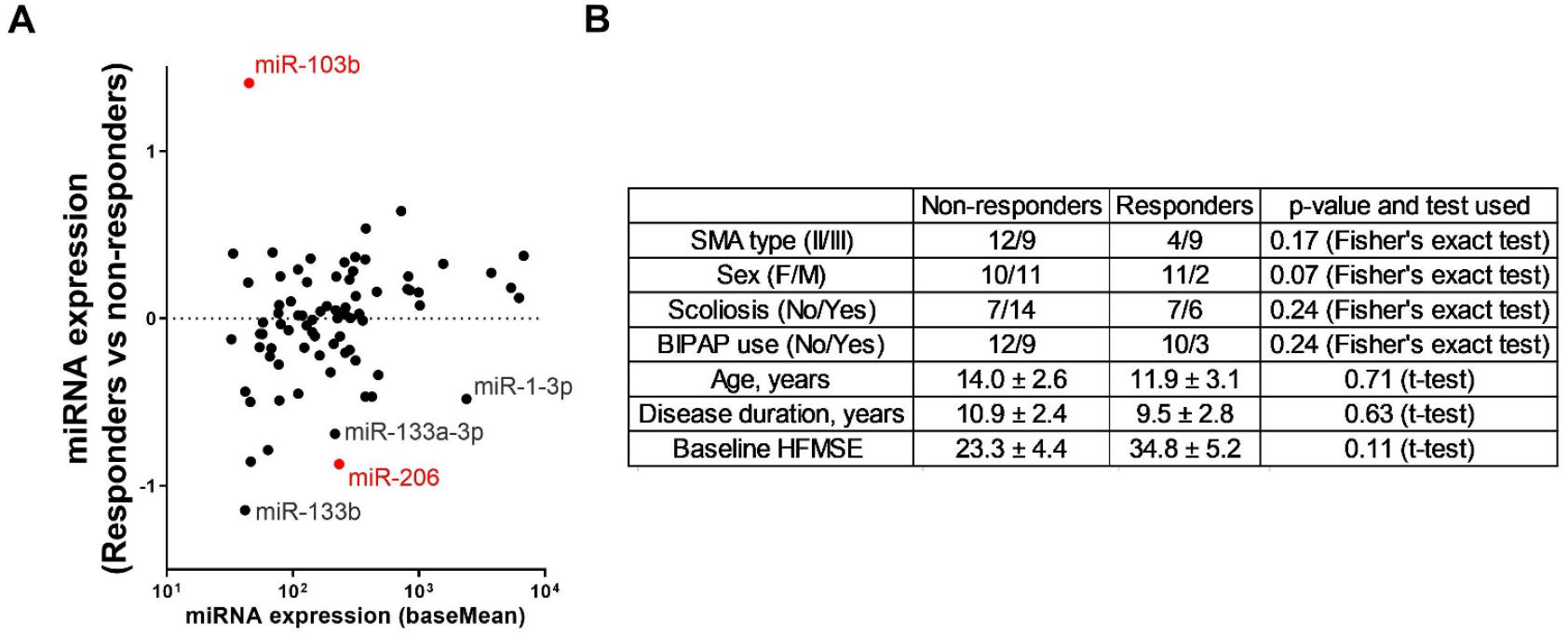
Pre-treatment miRNA signature predicts the response to nusinersen treatment. **(A)** MA plot of differential miRNA expression in responders (N=13, patients with HFMSE change ≥3 after 6 months of treatment) versus non-responders (N=21, HFMSE change ≤0). Data from additional 11 patients with HFMSE change of a single or two units (1,2) were excluded. Log 2 transformed fold change (y-axis), against mean miRNA abundance (x-axis). Red: significantly changed miRNAs (p<0.05, Wald test). Higher miR-103b levels and lower muscle enriched miRNA miR-1-3p, miR-133a/b and miR-206 levels are observed in prospective nusinersen responders. **(B)** A summary of the clinical data in non-responders and responders, showing that they are not significantly different between the groups. Numerical data are presented as mean ± sem. Categorical data was analyzed for significant differences in sex, type, scoliosis and BIPAP use distribution by Fisher’s exact test, while continuous data was analyzed for differences by student’s t-test. Notably, baseline HFMSE did not significantly differ between non-responders and responders (p=0.11).

The levels of miR-206 were negative predictors of positive treatment response, quantified as the change in the HFMSE final score (Spearman rho = -0.43, 95% CI: -0.67 to -0.09, p = 0.01, **Figure 3A**), but were not correlated with baseline HFMSE score **(Figure 3B)**. In addition, an orthogonal logistic regression model supported that miR-206 levels correlated with clinical response to nusinersen therapy (*χ*^2^ = 7, p = 0.008, Akaike’s information criterion (AIC) = 42.2, **Figure 3C**). We then tested whether miR-206 levels can predict differential response, without considering clinical information about disease duration and SMA type (II/III). We found that miR-206 is predictive of differential response to nusinersen independent of other baseline characteristics (**Figure 3D, E**). Therefore, miR-206 is able to predict differential response between two patients with similar baseline characteristics.

**Figure 3.**
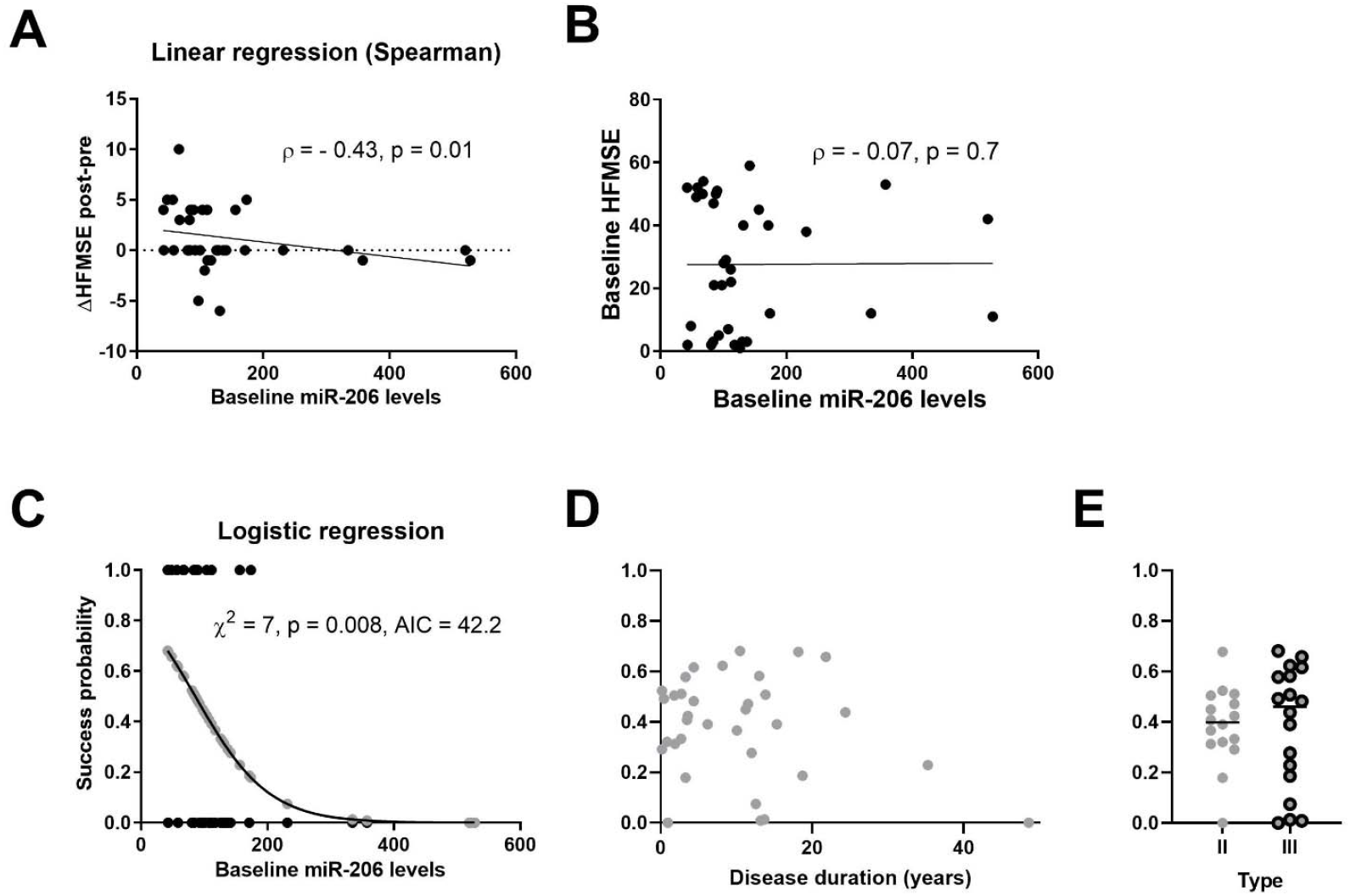
miR-206 levels predict the clinical improvement of SMA patients following 6-month nusinersen treatment. **(A)** Negative correlation of miR-206 levels with response to nusinersen therapy (as post-treatment – pre-treatment HFMSE score difference, Spearman rho = -0.43, p=0.01) **(B)** No correlation between miR-206 levels and HFMSE score at baseline **(C)** Logistic regression of baseline miR-206 levels on clinical dichotomized response to therapy: responders (N=13, HFMSE change ≥3 after 6 months of treatment) versus non-responders (N=21, HFMSE change ≤0). X-axis: miR-206 levels; Y-axis: probability of successful response predicted by a logistic regression model (depicted by grey dots on the curve). Black dots represent dichotomized clinical response (non-responders – 0, responders -1). Statistical significance of the logistic regression model goodness of fit was assessed by the chi-square test. AIC, Akaike’s information criterion. Success probabilities (Y-values the same as for grey dots in panel C) predicted by miR-206 **(D)** plotted against disease duration at treatment onset or **(E)** stratified by SMA type.

We next sought to validate the model on newly introduced data. We trained a machine learning procedure on 33 samples and tested the prediction on the single sample that was left out. The ‘leave one out cross validation’ (LOOCV) procedure, was reiterated 34 times. This analysis reveals that miR-206 predicts response more than can be expected at random (area under the receiver operating characteristic curve (ROC AUC) = 0.7, 95% CI: 0.51 – 0.88, p-value 0.06, **Figure S4**). As may be expected, the inclusion of eleven patients with intermediate clinical change, decreased the association between miR-206 levels and response to nusinersen, and reduced the accuracy of response prediction (**Figure S5**). None of the other myomiRs (miR-1-3p, miR-133a-3p or miR-133b, **Figure S6**) displayed a significant correlation with response to therapy. However, miR-103b levels correlated to the change in HFMSE score after therapy (**Figure S7**). Taken together, these data suggest that myomiRs predict the response to nusinersen therapy, and the miR-206 levels at baseline may predict the response to the nusinersen therapy.

### Multi-Feature Classifiers of Response to Nusinersen Therapy

We next considered simultaneously more than a single miRNA. The correlation of miR-206 and miR-133a-3p to clinical improvement and classification, when both are considered explanatory variables with weighted contributions to the model (multinomial logistic regression, equation available in supplementary table 1), was higher than when miR-206 was considered as a single feature (*χ*^2^ = 9, p = 0.01, AIC = 42.1; ROC AUC 0.74, 95% CI: 0.55 – 0.92, p = 0.022, **Figure 4A**). Similar to miR-206, miR-133a-3p did not correlate with baseline HFMSE score. None of the other miRNAs (miR-1-3p, miR-103b or miR-133b) displayed an improved prediction capacity when added on to miR-206 (**Figure S8A-C**). Thus, a feature composed of miR-206 and miR-133a-3p levels has an improved prediction capacity for nusinersen therapy response. Adding SMA type (III vs II) improved the correlation even further (<miR-206 + miR-133a-3p + SMA type>, *χ*^2^ = 12, p = 0.007, AIC = 41.35). We preserved the prediction capacity also by performing leave one out cross validation (LOOCV) (AUC = 0.73, 95% CI: p = 0.024, **Figure 4B**). Input about patient sex further improved prediction (<miR-206 + miR-133a-3p + SMA type+sex>, *χ*^2^ = 14.5, p = 0.006, AIC = 40.7; ROC AUC = 0.76, 95 CI%: 0.56 – 0.95, p = 0.01, **Figure 4C**), with males being less likely to respond to therapy. Patient age at baseline did not improve prediction (*χ*^2^ = 14.9, p = 0.01, AIC = 42.4; ROC AUC = 0.7, 95% CI: 0.475 – 0.925, p > 0.05, not shown), and was neither correlated with miR-133a (Spearman ρ = -0.01, p=0.96) nor miR-206 levels (Spearman ρ = 0.175, p=0.32), suggesting that miRNA levels predict response independently of age.

**Figure 4.**
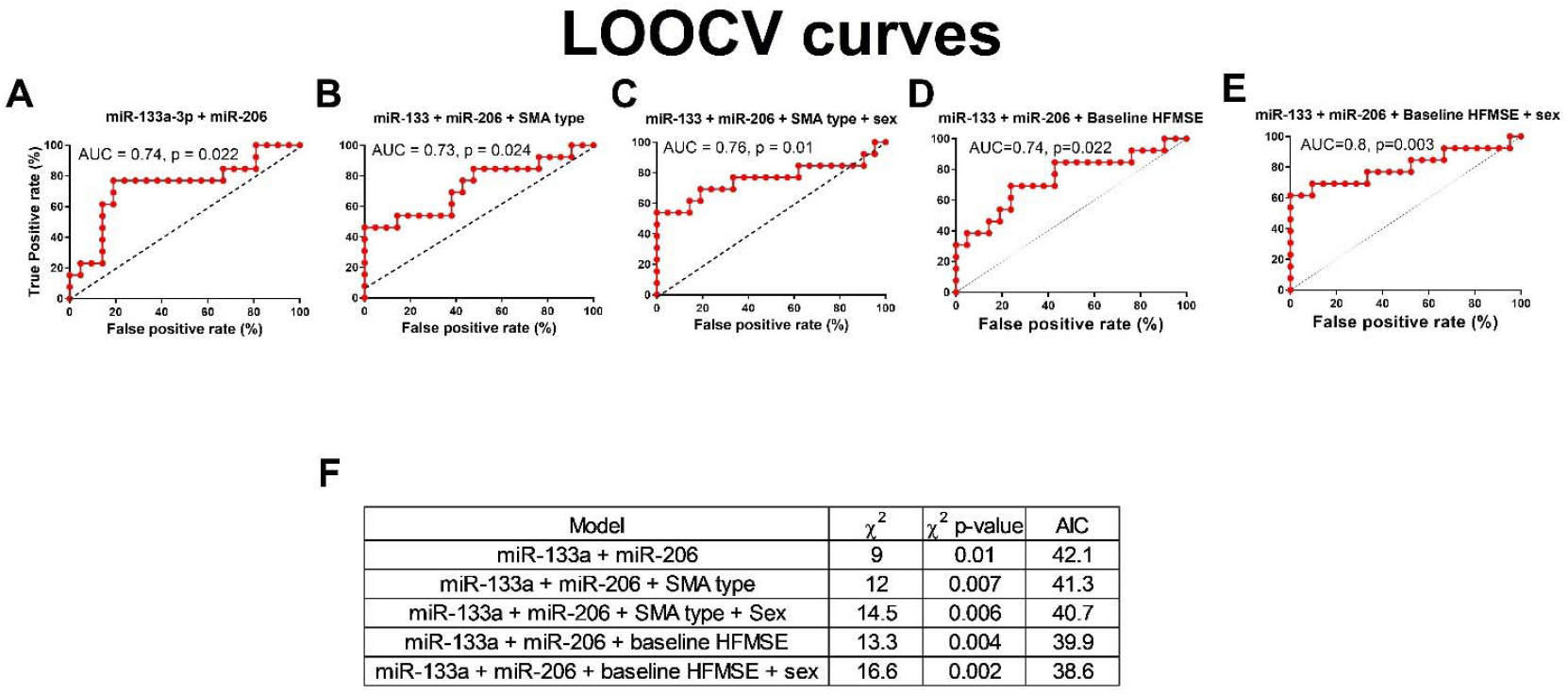
A predictor, based on miR-133a and miR-206 predicts response to nusinersen. ROC curves with leave one out cross validation for **(A)** miR-133a-3p + miR-206 **(B)** miR-133a-3p + miR-206 + SMA type **(C)** miR-133a-3p + miR-206 + SMA type + sex **(D)** miR-133a-3p + miR-206 + Baseline HFMSE score **(E)** miR-133a-3p + miR-206 + Baseline HFMSE score + sex. **(F)** A summary of logistic regression metrics for five multiple feature models.

Considering baseline HFMSE as a feature, together with miRNAs (miR-206 and miR-133a), yielded comparable, or slightly superior predictive capacities (without sex: *χ*^2^ = 12.1, p = 0.007, AIC = 41.2; ROC AUC 0.74, 95% CI: 0.55 – 0.92, p = 0.022, **Figure 4D;** with sex: *χ*^2^ = 15.2, p = 0.004, AIC = 40; ROC AUC 0.766, 95% CI: 0.58 – 0.95, p = 0.01, **Figure 4E**,**F**).

In addition to the multinomial regression of miR-133a and miR-206, we also devised a feature by normalization of the miRNA values by min-max scaling and summation of the scaled values. The new predictor, miR-133a/206, displays a *χ*^2^ of 9 (p = 0.003, **Figure 5A**), with a drop in predicted model errors relative to when miR-133a-3p and miR-206 are independently considered (AIC 40.2 vs. 42.1, respectively). The single standardized feature, miR-133a/206, displays ROC AUC of 0.74 (95% CI: 0.55 – 0.92, p = 0.02, **Figure 5B**) and information about SMA type (II or III based on clinical manifestation) and sex further improved correlation to clinical response and predictive capacity (*χ*^2^ = 14.5, p = 0.002, AIC = 38.8; ROC AUC = 0.784, 95% CI: 0.6 – 0.97, p = 0.006, **Figure 5C**). A model, which includes baseline HFMSE performs comparably (*χ*^2^ = 15.2, p = 0.0017, AIC = 38.0; ROC AUC = 0.78, 95% CI: 0.6 – 0.96, p = 0.007, **Figure 5D, E**).

**Figure 5.**
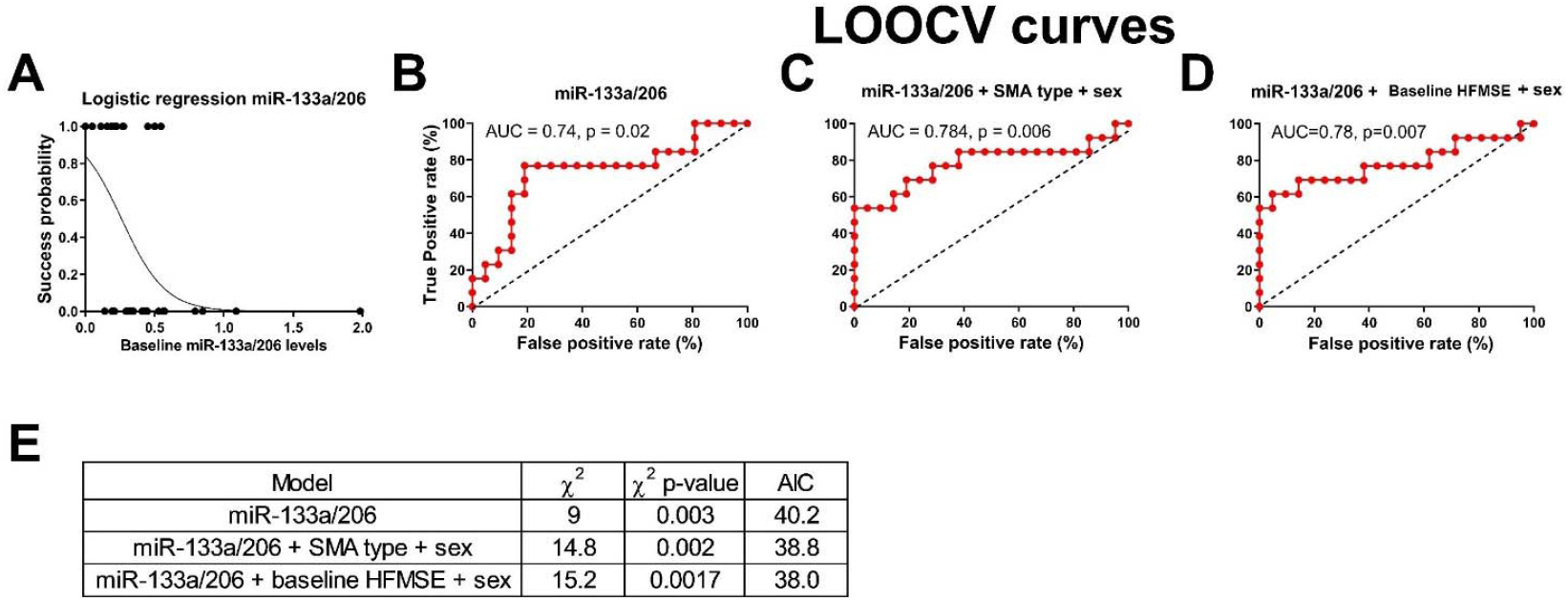
A predictor, based on a summation of the scaled values of miR-133a and miR-206 predicts response to nusinersen. **(A)** Logistic regression analysis of miR-133a/206 at baseline and clinical response to therapy after 6 months of nusinersen treatment. When the extreme sample whose miR-133a/206 score was 2 was excluded, chi square slightly decreases (from 9 to 8) and the p-values marginally increases (from 0.003 to 0.0046), suggesting the overall data and interpretation remain unchanged. ROC curves based on LOOCV for **(B)** miR-133a/206 **(C)** miR-133a/206 with SMA type and sex **(D)** miR-133/206 with baseline HFMSE and sex. **(E)** A summary of logistic regression metrics for three models.

Given inclusion of additional 11 patients with intermediate degree of clinical response to nusinersen therapy, miR-133a and miR-206 remained valuable predictors alone or in combination, with a notable reduction in accuracy (**Figure S9**).

Data was available for only 26 patients at longer follow up period of 12 months. Logistic regression analysis on miR-206 levels or on scaled levels of miR-133a/ miR-206 are consistent with earlier-time point analysis indicating that extremely high levels of myomiRs predict poor response to nusinersen (**Figure S10**). In summary, two miRNAs in the CSF are able to predict response to nusinersen therapy in SMA type II and III patients and may be useful for pre-treatment clinical decisions.

## Discussion

Given the expanding treatment options available for motor neuron diseases, the discovery of novel biomarkers to help track disease progression and therapeutic response is urgent. Here, we report novel findings derived from the first screening for small RNAs in the CSF of type II/III SMA patients treated with nusinersen. These data demonstrate that baseline levels of the myomiRs miR-133a and miR-206 predict the response to nusinersen therapy, suggesting that these myomiRs could play an important role in the clinical setting to help identify those patients most likely to demonstrate a more robust response to nusinersen therapy. In presenting these data, we hope to encourage incorporation of examination of CSF levels of miR-133a-3p and miR-206 as well as other myomiRs in longitudinal follow-up across cohorts of SMA patients receiving nusinersen or other novel therapies to help validate the current findings.

Identifying biomarkers to track SMA progression and the response to molecular therapies is an important focus of our group and others. We have previously demonstrated the usefulness of electrophysiologic measures of peripheral denervation to track peripheral motor unit integrity (39, 40). Circulating SMN protein levels is a candidate biomarker for monitoring SMA disease progression, although changes in peripheral SMN levels are not expected with at least two of the three currently FDA-approved therapies (41). CSF and plasma neurofilaments are markers of neuronal damage and have been identified as a potential treatment responsive biomarker in pediatric SMA and other neurodegenerative diseases, but baseline CSF or circulating neurofilament levels do not seem to necessarily predict treatment response in pediatric SMA patients (42-45). With three FDA-approved therapies and the anticipation of additional emerging treatments for SMA, comprehensive longitudinal cohort studies remain critically necessary to identify useful biomarkers that could predict the response to individual therapies.

One striking finding from our observations here is that myomiR levels were detected in the CSF and that CSF levels decreased in SMA patients that responded to nusinersen, when compared to those who presented a limited response. Although the myomiRs discussed here were identified via an unbiased screening approach in CSF, our data corroborate recent findings demonstrating that serum myomiRs levels are readouts of nusinersen response in SMA (25); We propose a model, whereby the presence of myomiRs in SMA CSF could be indicating blood-CSF barrier or blood-spinal cord disruption (46, 47), enabling the entrance of miRNAs that are derived from breakdown of muscle cells in the setting of acute denervation (22-24). Thus, lower myomiR levels prior to treatment with nusinersen, which predicts those with a better response potential, could reflect reduced muscle cell breakdown presumably associated with higher muscle mass and higher numbers of function motor units. In addition, a disease-associated damage to CSF barrier integrity might allow miRNA spilled from muscle to arrive at the CSF.

Another hypothesis is that miR-133a-3p and miR-206 may be involved as pathogenic factors, or may negatively modulate response to therapy. Thus, although not a prevailing hypothesis, the idea that miR-133a / miR-206 inhibitors could be of potential value in reducing disease progression, could be tested.

We previously demonstrated that serum creatinine, a product of skeletal muscle creatine metabolism, was also identified as a candidate biomarker to track SMA disease progression (48), and that creatinine maybe a sensitive predictor for the onset of denervation in infants with 3 *SMN2* copies (48). Similarly, circulating creatine kinase (CK) levels correlate with SMA severity and decreased during nusinersen therapy (49). Therefore, in the future creatine capacity to predict response to Nusinersen therapy may be tested by protein biomarkers, on their own, or in combination with miRNAs, such as miR-206/133a-3p.

Limitations of our study include a relatively small cohort and a limited follow up period. When larger cohorts are studied, over longer follow up periods, the use of RULM and 6MWT clinical scores in addition to HFMSE, may enable better clinical estimation. More work is required on additional cohorts and in creating means for quantification of absolute miRNA concentrations, before these findings can be applied to the clinic.

In summary, the current study demonstrates that the levels of muscle microRNAs miR-206 and miR-133a in CSF can predict the clinical response to nusinersen treatment in SMA patients. These novel findings have high clinical relevance and may prove useful in helping to design appropriate protocols intended to refine and improve treatment outcomes. Targeted approaches for quantification of miR-133a and miR-206 may increase in the future the accessibility to miRNA measurements and as a result, CSF miRNAs may be used as means to determine whether or not to initiate treatment. However, to get to that point, future studies should be performed in a bigger, ethnically-diverse, groups of patients and ethical issues should be considered.

## Data Availability

All data referred to in this manuscript are available on demand

## Data Availability Statement

The data that support the findings of this study are available from the corresponding author upon reasonable request.

## Acknowledgments

EH is the Mondry Family Professorial Chair and Head of the Andi and Larry Wolfe center for neuroimmunology and neuromodulation. Research at Hornstein lab is supported by CReATe consortium and ALSA (program: ‘Prognostic value of miRNAs in biofluids from ALS patients’), RADALA Foundation; AFM Telethon (20576); Weizmann - Brazil Center for Research on Neurodegeneration at Weizmann Institute of Science; the Minerva Foundation with funding from the Federal German Ministry for Education and Research, ISF Legacy Heritage Fund 828/17; Israel Science Foundation 135/16; 3497/21; 424/22, 425/22. Target ALS 118945; Thierry Latran Foundation for ALS Research. the European Research Council under the European Union’s Seventh Framework Programme (FP7/2007–2013)/ERC grant agreement number 617351; ERA-Net for Research Programmes on Rare Diseases (eRARE FP7) via Israel Ministry of Health; Dr. Sydney Brenner and friends, Edward and Janie Moravitz, A. Alfred Taubman through IsrALS, Yeda-Sela, Yeda-CEO, Israel Ministry of Trade and Industry; Y. Leon Benoziyo Institute for Molecular Medicine, Kekst Family Institute for Medical Genetics; David and Fela Shapell Family Center for Genetic Disorders Research; Crown Human Genome Center; Nathan, Shirley, Philip and Charlene Vener New Scientist Fund; Julius and Ray Charlestein Foundation; Fraida Foundation; Wolfson Family Charitable Trust; Adelis Foundation; Merck (United Kingdom); M. Halphen; and Estates of F. Sherr, L. Asseof, L. Fulop; Redhill Foundation – Sam and Jean Rothberg Charitable Trust. NSY was supported by the Israeli Council for Higher Education (CHE) via the Weizmann Data Science Research Center. IM was supported by Teva Pharmaceutical Industries Ltd. as part of the Israeli National Network of Excellence in Neuroscience (NNE, fellowship 117941). CRRA was funded by MGH Executive Committee on Research. KJS was funded by NIH NICHD R01HD054599, NIH NINDS R21NS108015, Biogen, AveXis and Cure SMA. We are grateful to all the patients and families who participated in this study.

## Supplementary tables and figures

**Supplementary table 1.** Logistic regression equations for the contribution of specific miRNAs clinical features or combinations thereof, to a prediction model. *1 is female, 2 is male. All equations refer to clinical response after 6 months unless noted otherwise.

| Model | Equation |
| --- | --- |
| miR-206 | $p = \frac{1}{1 + e^{-1.4868 + 0.0173 * miR-206}}$ |
| miR-133a + miR-206 | $p = \frac{1}{1 + e^{-2.22 + 0.02 * miR-133a + 0.01 * miR-206}}$ |
| miR-133a + miR-206 + SMA type | $p = \frac{1}{1 + e^{1.186 + 0.024 * miR-133a + 0.013 * miR-206 - 1.44 * type}}$ |
| miR-133a + miR-206 + SMA type + sex | $p = \frac{1}{1 + e^{0.11 + 0.02 * miR-133a + 0.01 * miR-206 - 1.66 * type + 1.53 * sex}}$ |
| miR-133a + miR-206 + HFMSE | $p = \frac{1}{1 + e^{1.45 + 0.024 * miR-133a + 0.013 * miR-206 - 0.36 * HFMSE}}$ |
| miR-133a + miR-206 + HFMSE + sex | $p = \frac{1}{1 + e^{3.2 + 0.02 * miR-133a + 0.01 * miR-206 - 0.05 * HFMSE + 1.77 * sex}}$ |
| miR-133a/206 | $p = \frac{1}{1 + e^{-1.67 + 6.35 * miR-133a/206}}$ |
| miR-133a/206 + SMA type + sex* | $p = \frac{1}{1 + e^{0.63 + 5.79 * miR-133a/206 - 1.66 * type + 1.54 * sex}}$ |
| miR-133a/206 + HFMSE + sex* | $p = \frac{1}{1 + e^{-2.64 + 6.15 * miR-133a/206 - 0.05 * HFMSE + 1.77 * sex}}$ |
| miR-1-3p | $p = \frac{1}{1 + e^{-0.44 + 0.0016 * miR-1-3p}}$ |
| miR-133a-3p | $p = \frac{1}{1 + e^{-1.04 + 0.026 * miR-133a-3p}}$ |
| miR-133b | $p = \frac{1}{1 + e^{0.15 + 0.012 * miR-133b}}$ |
| miR-103b | $p = \frac{1}{1 + e^{0.92 - 0.01 * miR-103b}}$ |
| miR-103b + miR-206 | $p = \frac{1}{1 + e^{-0.67 - 0.012 * miR-103b + 0.014 * miR-206}}$ |
| miR-133b + miR-206 | $p = \frac{1}{1 + e^{-1.74 + 0.01 * miR-133b + 0.017 * miR-206}}$ |
| miR-1-3p + miR-206 | $p = \frac{1}{1 + e^{-1.65 + 0.0008 * miR-1-3p + 0.0146 * miR-206}}$ |
| miR-206 12 months | $p = \frac{1}{1 + e^{-2.25 + 0.016 * miR-206}}$ |
| miR-133a/206 12 months | $p = \frac{1}{1 + e^{-1.5 + 3.5 * miR-133a/206}}$ |

**Supplementary table 2.** Logistic regression equations for the contribution of specific miRNAs clinical features or combinations thereof, to a prediction model, when additional 11 patients with intermediate response are included. *1 is female, 2 is male.

| Model | Equation |
| --- | --- |
| **miR-206** | $p = \frac{1}{1 + e^{-1.62 + 0.0125 * miR-206}}$ |
| **miR-133a + miR-206** | $p = \frac{1}{1 + e^{-1.7 + 0.0035 * miR-133a + 0.01 * miR-206}}$ |
| **miR-133a + miR-206 + SMA type** | $p = \frac{1}{1 + e^{1.005 + 0.003 * miR-133a + 0.0134 * miR-206 - 1.16 * type}}$ |
| **miR-133a + miR-206 + SMA type + sex** | $p = \frac{1}{1 + e^{0.31 + 0.005 * miR-133a + 0.0126 * miR-206 - 1.12 * type + 0.4 * sex}}$ |
| **miR-133a/206** | $p = \frac{1}{1 + e^{-1.35 + 5.38 * miR-133a/206}}$ |
| **miR-133a/206 + SMA type + sex\*** | $p = \frac{1}{1 + e^{0.46 + 5.96 * miR-133a/206 - 1.08 * type + 0.52 * sex}}$ |

**Supplementary Figure 1.**
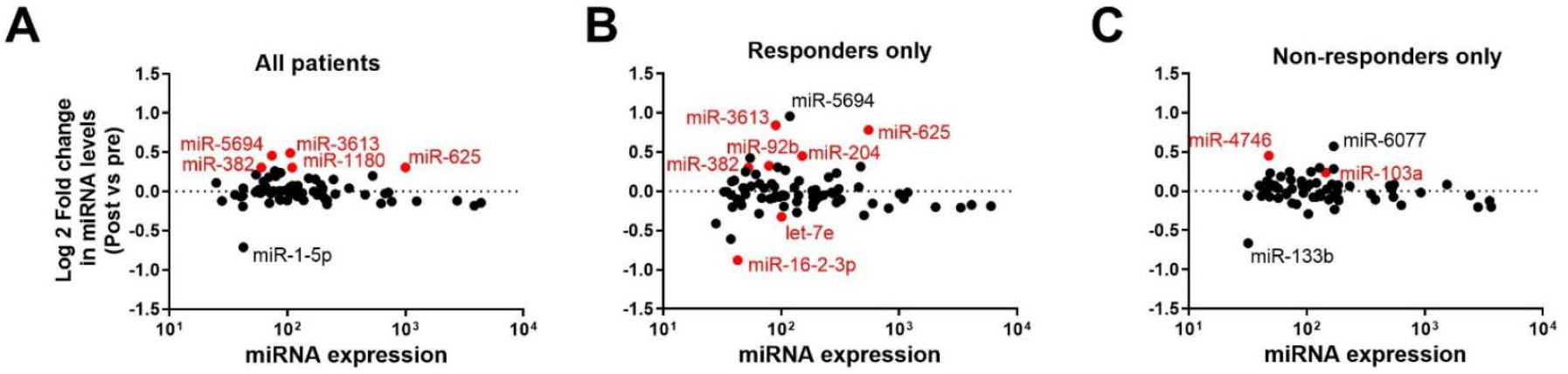
CSF miRNA signature after nusinersen treatment vs pre-treatment in the same patients. **(A)** MA plot showing, in the X-axis, miRNA levels averaged across CSF samples collected from 34 patients at both baseline (pre-treatment) and after 6 months of nusinersen treatment (68 samples in total). Y-axis, miRNA levels after treatment versus pre-treatment (log 2 of fold difference). Each dot represents a single miRNA gene. Dots above/below the horizontal dashed line are miRNAs with higher/lower levels post-treatment. MA plots showing of differential miRNA profile (at 6 months after therapy relative to baseline) of patients that **(B)** responded to nusinersen therapy (patients with change of HFMSE ≥ 3 points, 6 months after the molecular analysis of miRNAs), or **(C)** poorly responded (patients with change of HFMSE ≤ 0 points). Red dots represent miRNAs with p-value <0.05 (unadjusted) and log 2 fold change > 0.3 or <-0.3 (fold change > 1.2 or <0.8).

**Supplementary Figure 2.**
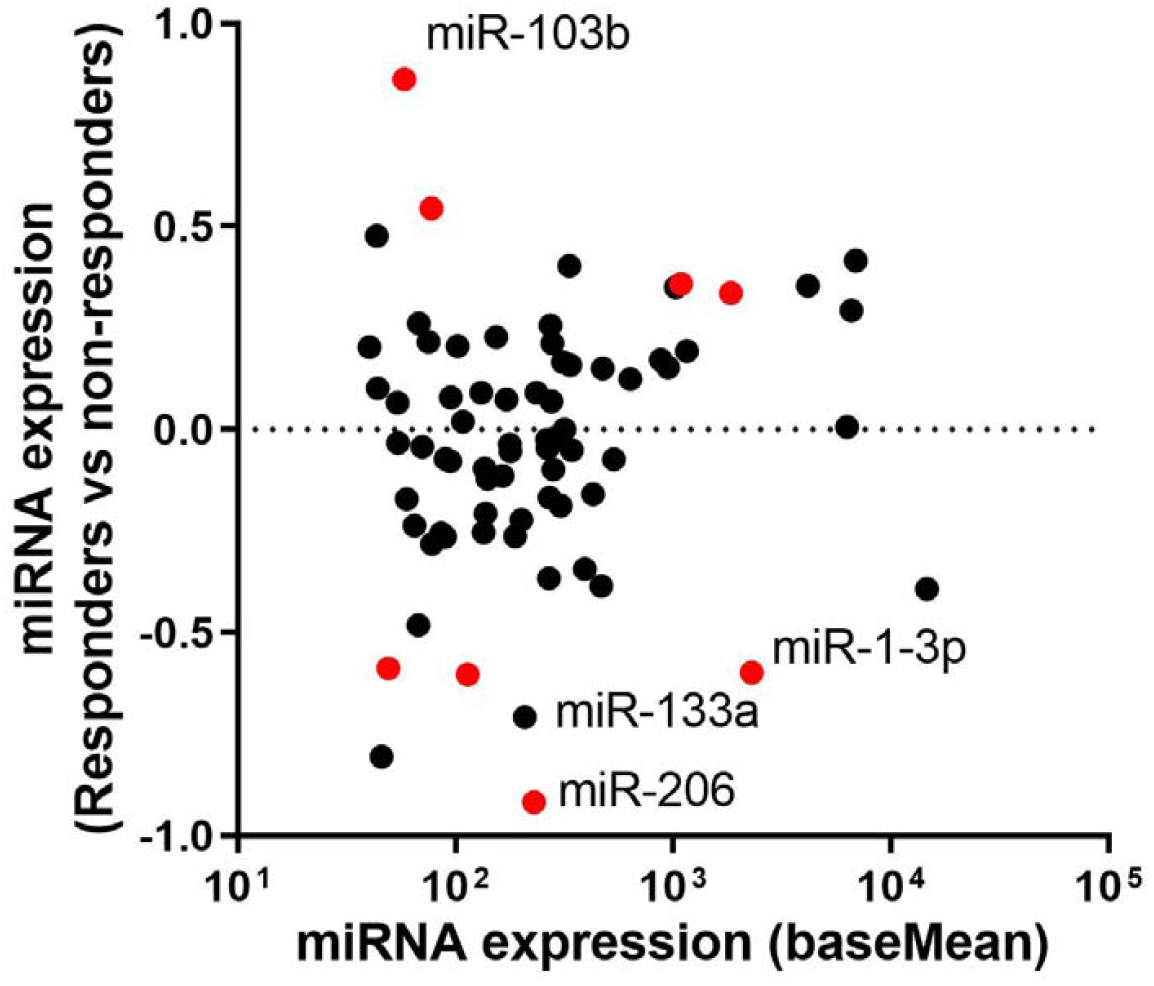
Pre-treatment miRNA signature predicts the response to nusinersen treatment in all 45 patients. MA plot of differential miRNA expression in responders (N=24, patients with HFMSE change ≥0 after 6 months of treatment) versus non-responders (N=21, HFMSE change ≤0). Log 2 transformed fold change (y-axis), against mean miRNA abundance (x-axis). Red: significantly changed miRNAs (p<0.05, Wald test).

**Supplementary Figure 3.**
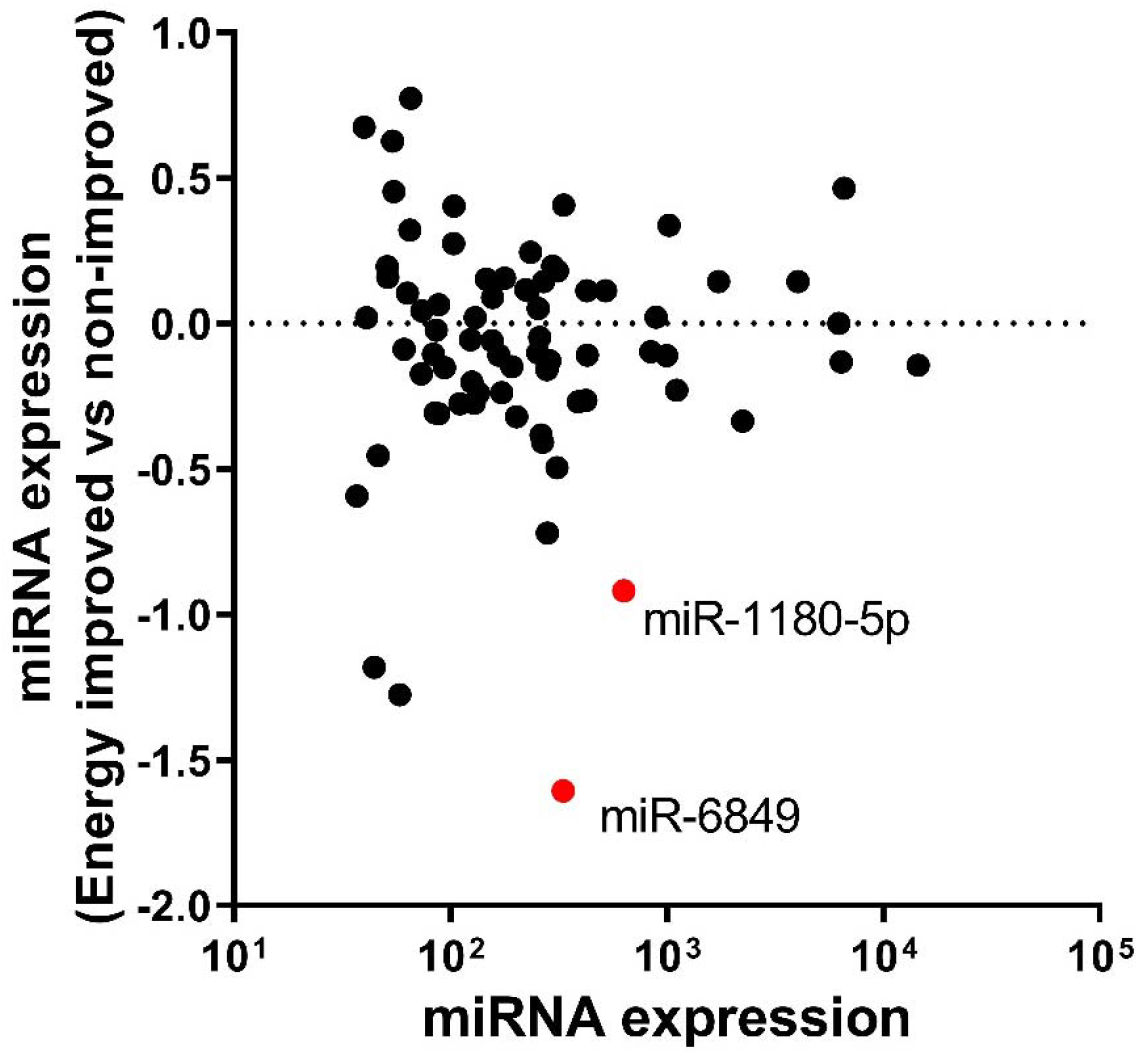
Pre-treatment miRNA signature in patients with subjective improvement of energy. MA plot of differential miRNA expression in patients reporting subjective energy improvement (N=29) vs those who did not report improvement (N=15). Log 2 transformed fold change (y-axis), against mean miRNA abundance (x-axis). Red: significantly changed miRNAs (p<0.05, Wald test).

**Supplementary Figure 4.**
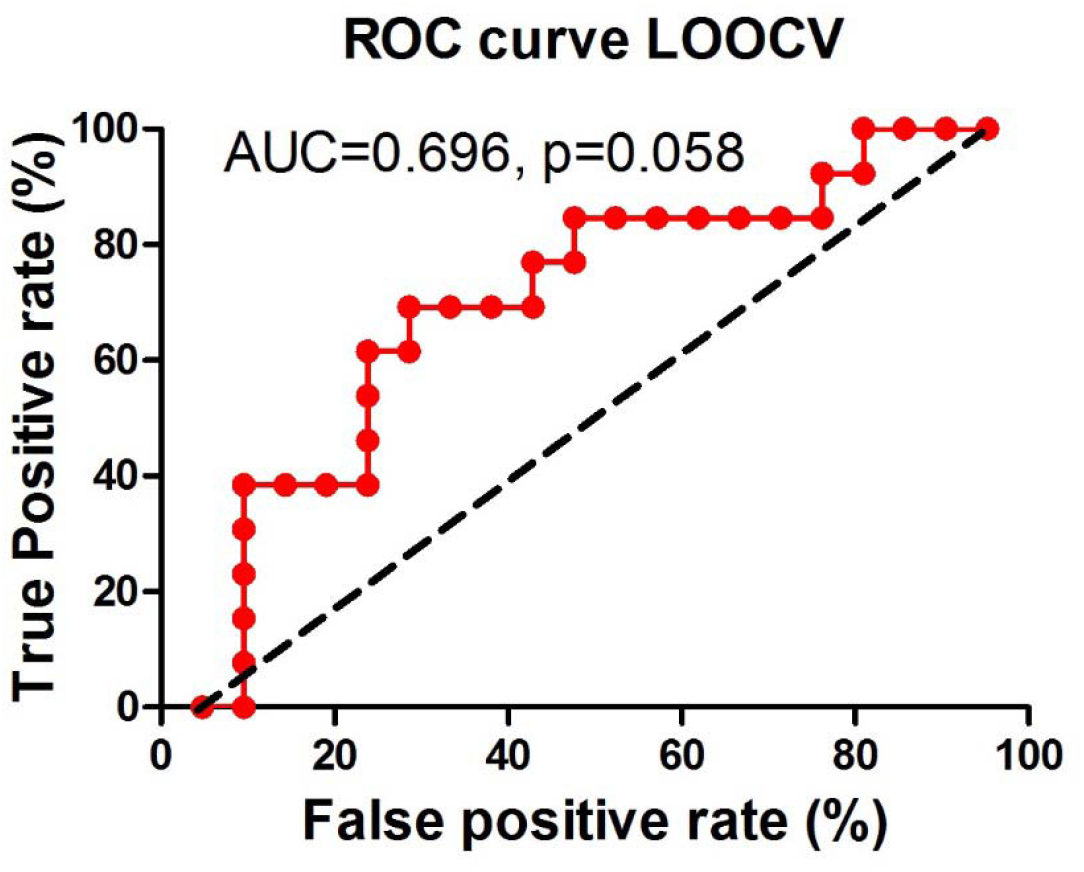
Receiver operating characteristic (ROC) curve based on miR-206 logistic regression and leave-one-out cross validation (LOOCV). True positive rate (sensitivity, y-axis) against false positive rate (100%-specificity, x-axis). Values of predicted success probability in each patient, as a cut-off for binary classification.

**Supplementary Figure 5.**
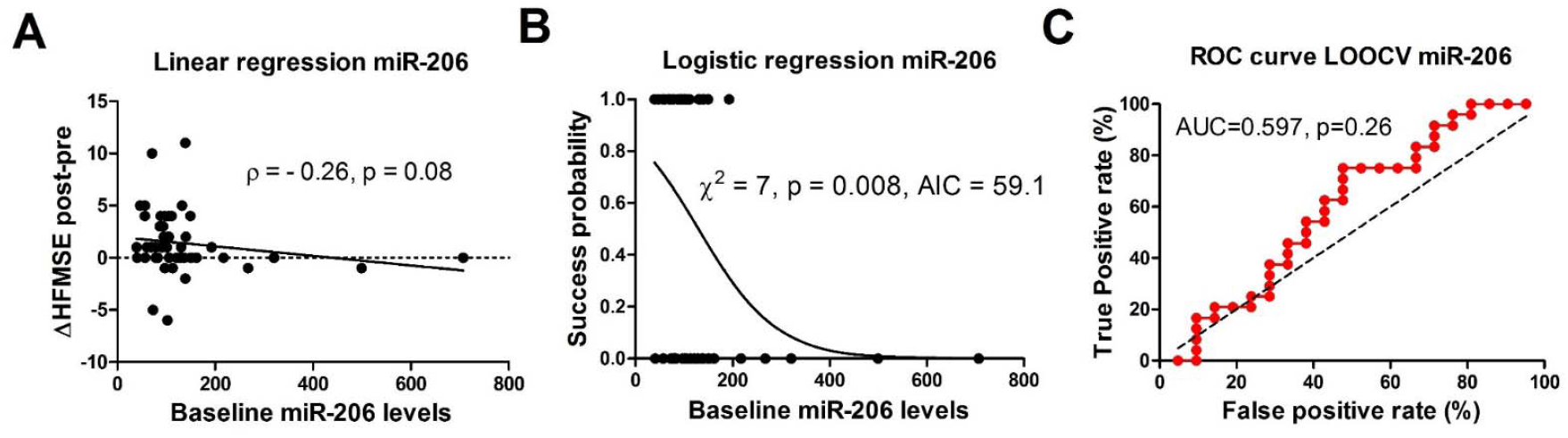
miR-206 levels predict clinical improvement of SMA patients following 6-month nusinersen treatment in all 45 patients. **(A)** Negative correlation of miR-206 levels with response to nusinersen therapy (as post-treatment – pre-treatment HFMSE score difference, Spearman rho = -0.28, p=0.08) **(B)** Logistic regression of baseline miR-206 levels on clinical dichotomized response to therapy: responders (N=24, HFMSE change ≥0 after 6 months of treatment) versus non-responders (N=21, HFMSE change ≤0). Statistical significance of the logistic regression model goodness of fit was assessed by the chi-square test. AIC, Akaike’s information criterion. **(C)** Receiver operating characteristic (ROC) curve based on miR-206 logistic regression and leave-one-out cross validation (LOOCV).

**Supplementary Figure 6.**
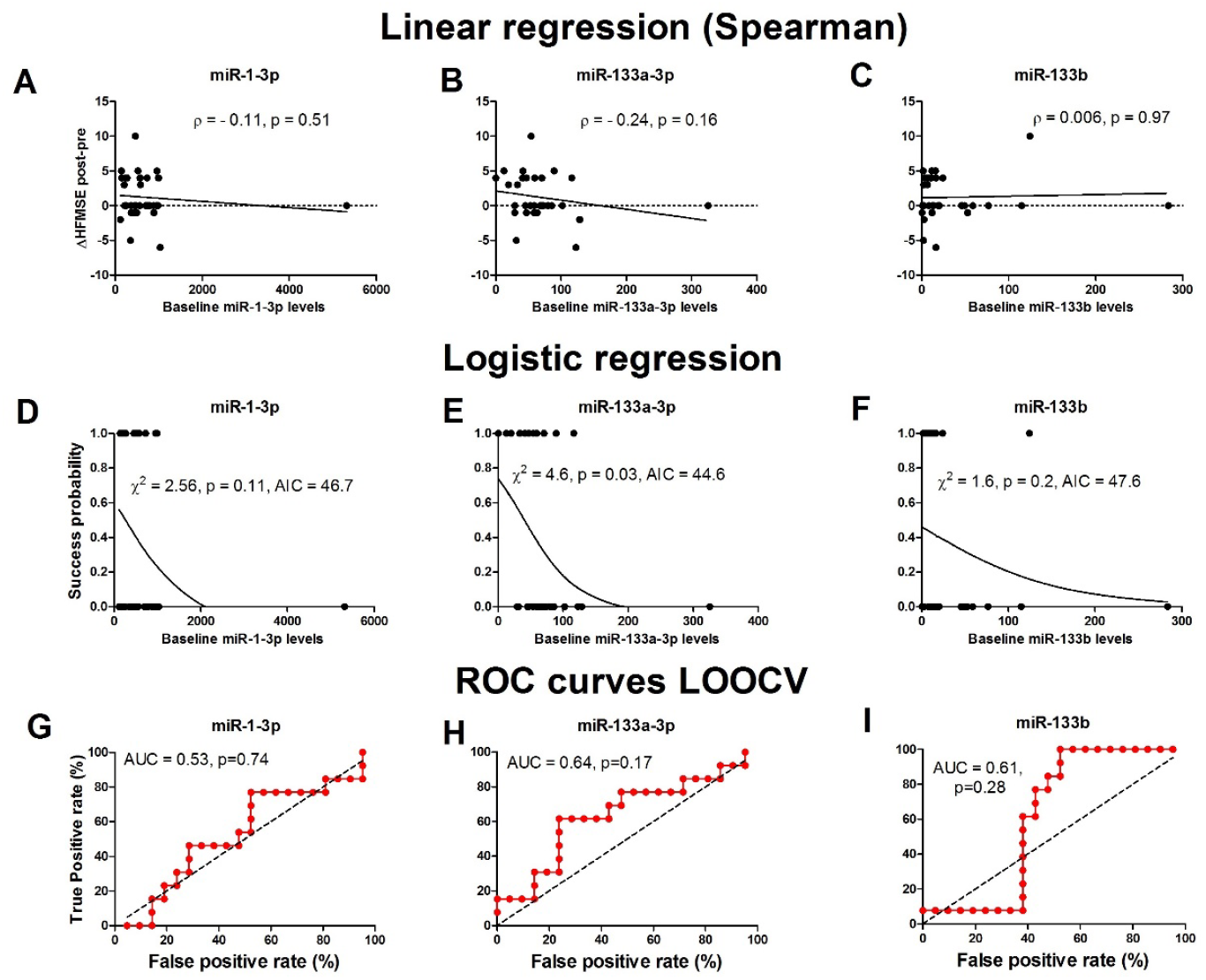
Association of myomiRs with clinical improvement of SMA patients during nusinersen treatment. **Spearman correlation** of baseline miR-1-3p **(A)**, miR-133a-3p **(B)** and miR-133b **(C)** with the difference in HFMSE score following nusinersen. **Logistic regression** analysis of baseline miR-1-3p **(D)**, miR-133a-3p **(E)** and miR-133b **(F)** levels on clinical response, after 6 months of nusinersen treatment. **ROC curves** based on LOOCV for miR-1-3p **(G)**, miR-133a-3p **(H)** and miR-133b **(I)**.

**Supplementary Figure 7.**
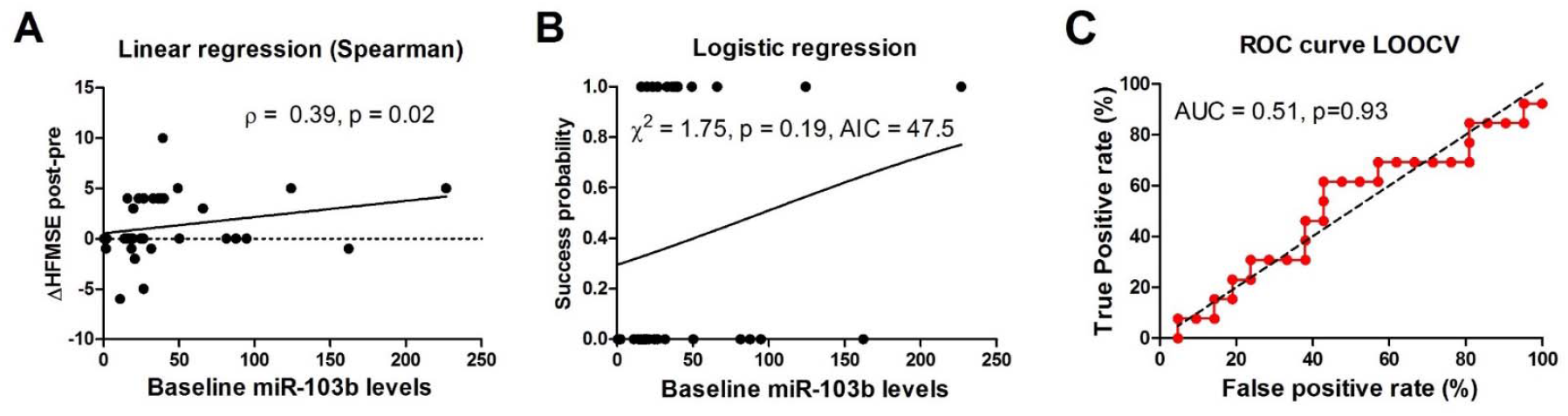
Increased miR-103b in the CSF modestly predicts clinical improvement of SMA patients during nusinersen treatment. **(A)** miR-103b levels at baseline exhibit a positive and significant correlation (Spearman rho = 0.39, p=0.02) with the difference in HFMSE score following nusinersen. **(B)** Logistic regression analysis of miR-103b levels at baseline and clinical response to therapy after 6 months of nusinersen treatment. **(C)** ROC curve based on LOOCV for the logistic regression model.

**Supplementary Figure 8.**
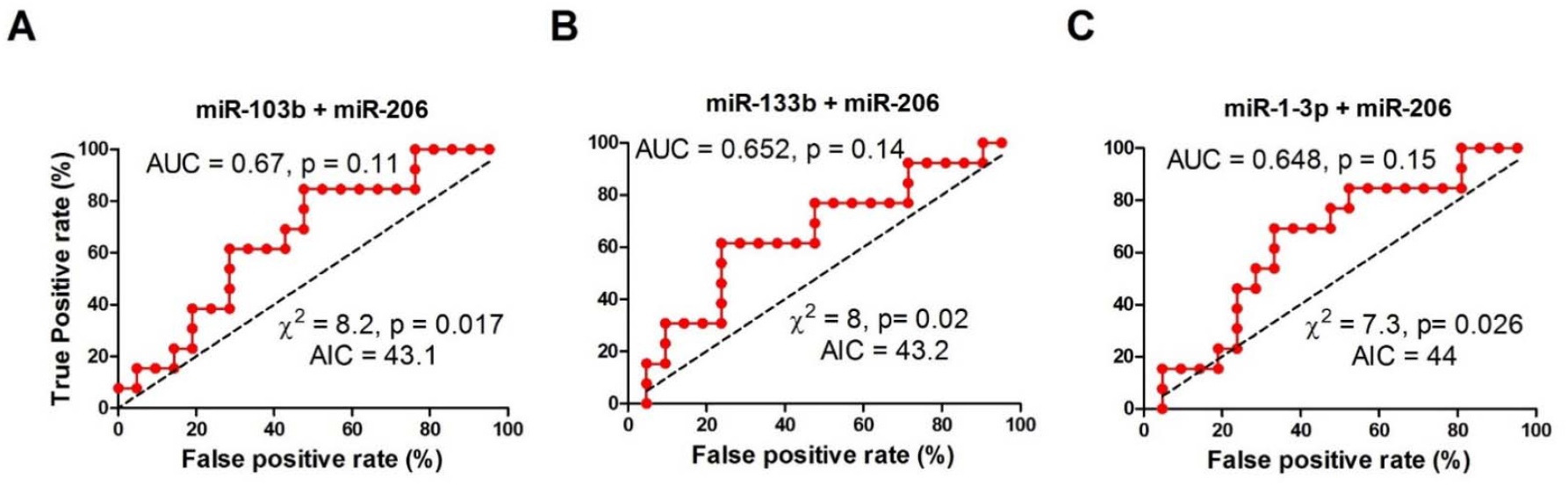
ROC curves for models of **(A)** miR-206 + miR-103b **(B)** miR-206 + miR-133b and **(C)** miR-206 + miR-1-3p.

**Supplementary Figure 9.**
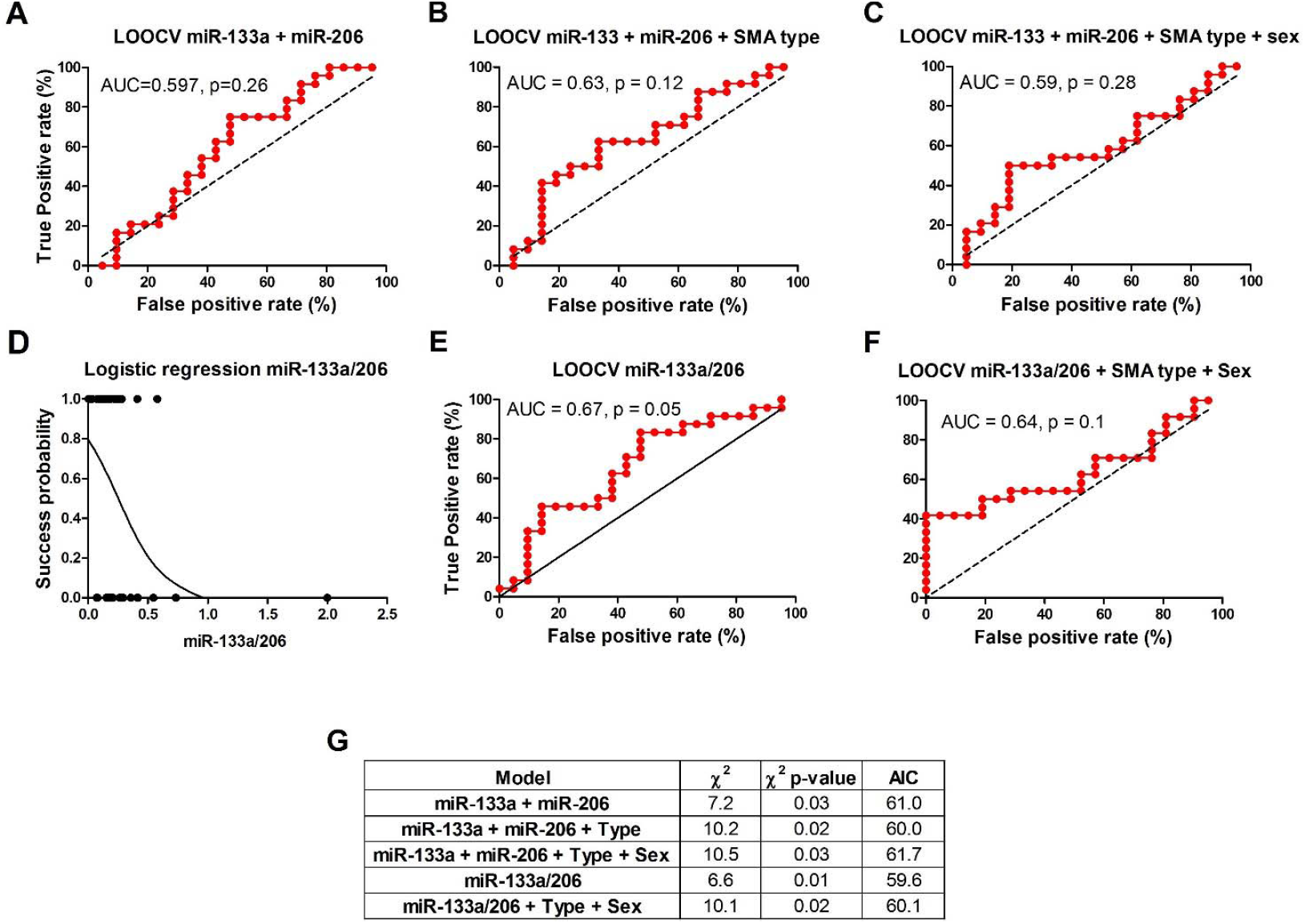
Prediction of response to nusinersen by combination of miR-133a, miR-206 and clinical features in all 45 patients. ROC curves **(A)** miR-133a-3p + miR-206 **(B)** miR-133a-3p + miR-206 + SMA type **(C)** miR-133a-3p + miR-206 + SMA type + sex with leave one out cross validation. **(D)** Logistic regression analysis of miR-133a/206 at baseline and clinical response to therapy after 6 months of nusinersen treatment. ROC curves based on LOOCV for **(E)** miR-133a/206 and **(F)** miR-133a/206 with SMA type and sex. **(G)** A summary of logistic regression parameters for five multiple feature models.

**Supplementary Figure 10.**
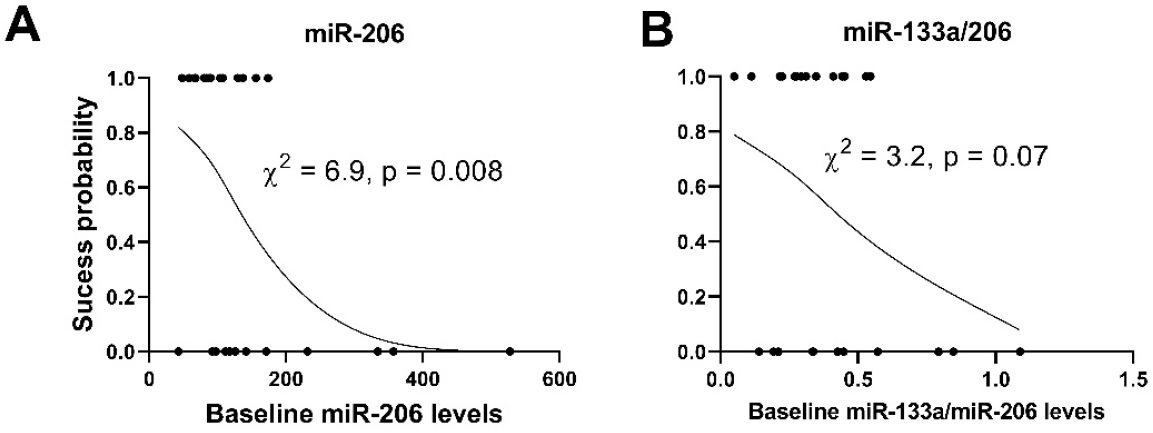
Prediction of response to nusinersen after 12 months. ROC curves Logistic regression analysis of miR-206 **(A)** and miR-133a/206 **(B)** at baseline and clinical response to therapy after 12 months of nusinersen treatment.

## References

1. Ogino S, Leonard DG, Rennert H, Wilson RB. Spinal muscular atrophy genetic testing experience at an academic medical center. J Mol Diagn. 2002;4(1):53–8.

2. Verhaart IEC, Robertson A, Wilson IJ, Aartsma-Rus A, Cameron S, Jones CC, et al. Prevalence, incidence and carrier frequency of 5q-linked spinal muscular atrophy - a literature review. Orphanet J Rare Dis. 2017;12(1):124.

3. Prior TW, Leach ME, Finanger E. Spinal Muscular Atrophy. In: Adam MP, Ardinger HH, Pagon RA, Wallace SE, Bean LJH, Mirzaa G, et al., editors. GeneReviews((R)). Seattle (WA)1993.

4. Brahe C, Servidei S, Zappata S, Ricci E, Tonali P, Neri G. Genetic homogeneity between childhood-onset and adult-onset autosomal recessive spinal muscular atrophy. Lancet. 1995;346(8977):741–2.

5. Clermont O, Burlet P, Lefebvre S, Burglen L, Munnich A, Melki J. SMN gene deletions in adult-onset spinal muscular atrophy. Lancet. 1995;346(8991-8992):1712–3.

6. Wadman RI, Stam M, Gijzen M, Lemmink HH, Snoeck IN, Wijngaarde CA, et al. Association of motor milestones, SMN2 copy and outcome in spinal muscular atrophy types 0-4. J Neurol Neurosurg Psychiatry. 2017;88(4):365–7.

7. Zerres K, Rudnik-Schoneborn S, Forrest E, Lusakowska A, Borkowska J, Hausmanowa-Petrusewicz I. A collaborative study on the natural history of childhood and juvenile onset proximal spinal muscular atrophy (type II and III SMA): 569 patients. J Neurol Sci. 1997;146(1):67–72.

8. Groen EJN, Talbot K, Gillingwater TH. Advances in therapy for spinal muscular atrophy: promises and challenges. Nat Rev Neurol. 2018;14(4):214–24.

9. Faravelli I, Nizzardo M, Comi GP, Corti S. Spinal muscular atrophy--recent therapeutic advances for an old challenge. Nat Rev Neurol. 2015;11(6):351–9.

10. Mailman MD, Heinz JW, Papp AC, Snyder PJ, Sedra MS, Wirth B, et al. Molecular analysis of spinal muscular atrophy and modification of the phenotype by SMN2. Genet Med. 2002;4(1):20–6.

11. Feldkotter M, Schwarzer V, Wirth R, Wienker TF, Wirth B. Quantitative analyses of SMN1 and SMN2 based on real-time lightCycler PCR: fast and highly reliable carrier testing and prediction of severity of spinal muscular atrophy. Am J Hum Genet. 2002;70(2):358–68.

12. Crawford TO, Paushkin SV, Kobayashi DT, Forrest SJ, Joyce CL, Finkel RS, et al. Evaluation of SMN protein, transcript, and copy number in the biomarkers for spinal muscular atrophy (BforSMA) clinical study. PLoS One. 2012;7(4):e33572.

13. FDA Approval of Nusinersen for Spinal Muscular Atrophy Makes 2016 the Year of Splice Modulating Oligonucleotides. Nucleic Acid Therapeutics. 2017;27(2):67–9.

14. Messina S. New Directions for SMA Therapy. Journal of Clinical Medicine. 2018;7(9):251.

15. Hagenacker T, Wurster CD, Gunther R, Schreiber-Katz O, Osmanovic A, Petri S, et al. Nusinersen in adults with 5q spinal muscular atrophy: a non-interventional, multicentre, observational cohort study. Lancet Neurol. 2020;19(4):317–25.

16. Walter MC, Wenninger S, Thiele S, Stauber J, Hiebeler M, Greckl E, et al. Safety and Treatment Effects of Nusinersen in Longstanding Adult 5q-SMA Type 3 - A Prospective Observational Study. J Neuromuscul Dis. 2019;6(4):453–65.

17. Emde A, Eitan C, Liou LL, Libby RT, Rivkin N, Magen I, et al. Dysregulated miRNA biogenesis downstream of cellular stress and ALS-causing mutations: a new mechanism for ALS. EMBO J. 2015;34(21):2633–51.

18. Reichenstein I, Eitan C, Diaz-Garcia S, Haim G, Magen I, Siany A, et al. Human genetics and neuropathology suggest a link between miR-218 and amyotrophic lateral sclerosis pathophysiology. Science Translational Medicine. 2019;11(523):eaav5264.

19. Haramati S, Chapnik E, Sztainberg Y, Eilam R, Zwang R, Gershoni N, et al. miRNA malfunction causes spinal motor neuron disease. Proc Natl Acad Sci U S A. 2010;107(29):13111–6.

20. Chen J-F, Mandel EM, Thomson JM, Wu Q, Callis TE, Hammond SM, et al. The role of microRNA-1 and microRNA-133 in skeletal muscle proliferation and differentiation. Nature genetics. 2006;38(2):228–33.

21. Chen J-F, Tao Y, Li J, Deng Z, Yan Z, Xiao X, et al. microRNA-1 and microRNA-206 regulate skeletal muscle satellite cell proliferation and differentiation by repressing Pax7. Journal of Cell Biology. 2010;190(5):867–79.

22. de Andrade HM, de Albuquerque M, Avansini SH, de SRC, Dogini DB, Nucci A, et al. MicroRNAs-424 and 206 are potential prognostic markers in spinal onset amyotrophic lateral sclerosis. J Neurol Sci. 2016;368:19–24.

23. Toivonen JM, Manzano R, Olivan S, Zaragoza P, Garcia-Redondo A, Osta R. MicroRNA-206: a potential circulating biomarker candidate for amyotrophic lateral sclerosis. PLoS One. 2014;9(2):e89065.

24. Waller R, Goodall EF, Milo M, Cooper-Knock J, Da Costa M, Hobson E, et al. Serum miRNAs miR-206, 143-3p and 374b-5p as potential biomarkers for amyotrophic lateral sclerosis (ALS). Neurobiol Aging. 2017;55:123–31.

25. Bonanno S, Marcuzzo S, Malacarne C, Giagnorio E, Masson R, Zanin R, et al. Circulating MyomiRs as Potential Biomarkers to Monitor Response to Nusinersen in Pediatric SMA Patients. Biomedicines. 2020;8(2):21.

26. Coenen-Stass AML, Magen I, Brooks T, Ben-Dov IZ, Greensmith L, Hornstein E, et al. Evaluation of methodologies for microRNA biomarker detection by next generation sequencing. RNA Biol. 2018;15(8):1133–45.

27. Magen I, Yacovzada N-S, Warren JD, Heller C, Swift I, Bobeva Y, et al. miRNA biomarkers for diagnosis of ALS and FTD, developed by a nonlinear machine learning approach. medRxiv. 2021:2020.01.22.20018408.

28. Magen I, Yacovzada NS, Yanowski E, Coenen-Stass A, Grosskreutz J, Lu C-H, et al. Circulating miR-181 is a prognostic biomarker for amyotrophic lateral sclerosis. bioRxiv. 2021:833079.

29. Glanzman AM, Mazzone ES, Young SD, Gee R, Rose K, Mayhew A, et al. Evaluator Training and Reliability for SMA Global Nusinersen Trials1. J Neuromuscul Dis. 2018;5(2):159–66.

30. O’Hagen JM, Glanzman AM, McDermott MP, Ryan PA, Flickinger J, Quigley J, et al. An expanded version of the Hammersmith Functional Motor Scale for SMA II and III patients. Neuromuscul Disord. 2007;17(9-10):693–7.

31. Mercuri E, Darras BT, Chiriboga CA, Day JW, Campbell C, Connolly AM, et al. Nusinersen versus Sham Control in Later-Onset Spinal Muscular Atrophy. N Engl J Med. 2018;378(7):625–35.

32. Swoboda KJ, Scott CB, Reyna SP, Prior TW, LaSalle B, Sorenson SL, et al. Phase II open label study of valproic acid in spinal muscular atrophy. PLoS One. 2009;4(5):e5268.

33. Kohen R, Barlev J, Hornung G, Stelzer G, Feldmesser E, Kogan K, et al. UTAP: User-friendly Transcriptome Analysis Pipeline. BMC bioinformatics. 2019;24:154.

34. Kozomara A, Griffiths-Jones S. miRBase: annotating high confidence microRNAs using deep sequencing data. Nucleic Acids Res. 2014;42(Database issue):D68–73.

35. Love MI, Huber W, Anders S. Moderated estimation of fold change and dispersion for RNA-seq data with DESeq2. Genome Biol. 2014;15(12):550.

36. Multiple Logistic Regression. Applied Logistic Regression 2000. p. 31–46.

37. Akaike H. Information Theory and an Extension of the Maximum Likelihood Principle. In: Parzen E, Tanabe K, Kitagawa G, editors. Selected Papers of Hirotugu Akaike. New York, NY: Springer New York; 1998. p. 199–213.

38. Finkel RS, Mercuri E, Darras BT, Connolly AM, Kuntz NL, Kirschner J, et al. Nusinersen versus Sham Control in Infantile-Onset Spinal Muscular Atrophy. N Engl J Med. 2017;377(18):1723–32.

39. Bromberg MB, Swoboda KJ. Motor unit number estimation in infants and children with spinal muscular atrophy. Muscle Nerve. 2002;25(3):445–7.

40. Lewelt A, Krosschell KJ, Scott C, Sakonju A, Kissel JT, Crawford TO, et al. Compound muscle action potential and motor function in children with spinal muscular atrophy. Muscle Nerve. 2010;42(5):703–8.

41. Alves CRR, Zhang R, Johnstone AJ, Garner R, Eichelberger EJ, Lepez S, et al. Whole blood survival motor neuron protein levels correlate with severity of denervation in spinal muscular atrophy. Muscle Nerve. 2020;62(3):351–7.

42. Darras BT, Crawford TO, Finkel RS, Mercuri E, De Vivo DC, Oskoui M, et al. Neurofilament as a potential biomarker for spinal muscular atrophy. Ann Clin Transl Neurol. 2019;6(5):932–44.

43. Eichelberger EJ, Alves CRR, Zhang R, Petrillo M, Cullen P, Farwell W, et al. Increased systemic HSP70B levels in spinal muscular atrophy infants. Ann Clin Transl Neurol. 2021.

44. De Vivo DC, Bertini E, Swoboda KJ, Hwu WL, Crawford TO, Finkel RS, et al. Nusinersen initiated in infants during the presymptomatic stage of spinal muscular atrophy: Interim efficacy and safety results from the Phase 2 NURTURE study. Neuromuscul Disord. 2019;29(11):842–56.

45. Winter B, Guenther R, Ludolph AC, Hermann A, Otto M, Wurster CD. Neurofilaments and tau in CSF in an infant with SMA type 1 treated with nusinersen. J Neurol Neurosurg Psychiatry. 2019;90(9):1068–9.

46. Müschen LH, Osmanovic A, Binz C, Jendretzky KF, Ranxha G, Bronzlik P, et al. Cerebrospinal Fluid Parameters in Antisense Oligonucleotide-Treated Adult 5q-Spinal Muscular Atrophy Patients. Brain Sciences. 2021;11(3):296.

47. Somers E, Lees RD, Hoban K, Sleigh JN, Zhou H, Muntoni F, et al. Vascular Defects and Spinal Cord Hypoxia in Spinal Muscular Atrophy. Annals of Neurology. 2016;79(2):217–30.

48. Alves CRR, Zhang R, Johnstone AJ, Garner R, Nwe PH, Siranosian JJ, et al. Serum creatinine is a biomarker of progressive denervation in spinal muscular atrophy. Neurology. 2020;94(9):e921–e31.

49. Freigang M, Wurster CD, Hagenacker T, Stolte B, Weiler M, Kamm C, et al. Serum creatine kinase and creatinine in adult spinal muscular atrophy under nusinersen treatment. Ann Clin Transl Neurol. 2021.

